# Cross-cultural adaptation and validation of the “COVID Stress Scales” in Greek

**DOI:** 10.1101/2022.02.28.22271615

**Authors:** Petros Galanis, Irene Vraka, Aglaia Katsiroumpa, Olympia Konstantakopoulou, Olga Siskou, Eleftheria Zogaki, Daphne Kaitelidou

## Abstract

**Background:** During the COVID-19 pandemic, several instruments were developed to measure the psychological impact of COVID-19, such as fear, anxiety, post-traumatic stress, phobia, etc.

**Objective:** To adapt cross-cultural and validate the “COVID Stress Scales” in Greek.

**Methods:** We conducted a cross-sectional study with 200 participants between November 2021 to February 2022. All participants were adults, and a convenience sample was obtained. We applied the forward-backward translation method to create a Greek version of the “COVID Stress Scales”. We assessed reliability of the questionnaire with test-retest method in a 10-day window, and we assessed validity of the questionnaire with exploratory factor analysis.

**Results:** Our five-factor model explained 72% of the variance and totally confirmed the factors of the initial “COVID Stress Scales”. In particular, we found the following five factors: (a) COVID-19 danger and contamination (eleven items), (b) COVID-19 socioeconomic consequences (six items), (c) COVID-19 xenophobia (six items), (d) COVID-19 traumatic stress (six items), and (e) COVID-19 compulsive checking (six items). Cronbach’s coefficients alpha for the five factors that emerged from the exploratory factor analysis were greater than 0.89 indicating excellent internal reliability.

**Conclusions:** We found that the “COVID Stress Scales” is a reliable and valid tool to measure stress due to the COVID-19 in the Greek population.

## Introduction

Distress, anxiety, fear, and several other psychological symptoms are common during the COVID-19 pandemic (Bareeqa et al., 2021; Cénat et al., 2021; Galanis, Andreadaki, et al., 2021; Galanis, Petrogianni, et al., 2021; Patelarou et al., 2022; Yuan et al., 2021; Zhang et al., 2021). For instance, according to studies in China, more than 25% of the general population experienced moderate to severe levels of stress and anxiety during the COVID-19 pandemic (Qiu et al., 2020; Wang et al., 2020). The psychological management of the COVID-19 is essential since higher levels of perceived threat from the COVID-19 and fear of COVID-19 are associated with increased willingness to get vaccinated against COVID-19 (Galanis et al., 2022; Galanis, Vraka, et al., 2021; Patelarou et al., 2021). On the other hand, increased anxiety is related with socially disruptive behaviors, such as panic buying and surging unnecessarily into hospitals when people misinterpret their minor symptoms as signs of serious disease (Asmundson & Taylor, 2020a, 2020b).

Anxiety is important in shaping behavioral responses to the COVID-19 pandemic and COVID-19 stressors affect almost everyone in the world to different extent. During the COVID-19 pandemic, several instruments were developed to measure the psychological impact of COVID-19, such as fear, anxiety, post-traumatic stress, phobia, etc. (Ahorsu et al., 2020; Ahuja, 2021; Arpaci et al., 2020; Forte et al., 2020; Geldsetzer, 2020; Kira et al., 2021; Mertens et al., 2020; Pérez-Fuentes et al., 2020; Petzold et al., 2020; Varshney et al., 2020; Zhong et al., 2020). Many scales have a unidimensional structure, while others consist of factors, such as trauma, phobia, post-traumatic stress, anxiety, fear, etc. Moreover, construct and content validity of some scales is not evaluated, thus limiting the validity of these tools.

Among others, the “COVID Stress Scales” are one of the first scales worldwide that are developed to assess COVID-19 related stress (Taylor et al., 2020). The aim of our study was to adapt cross-cultural and validate the “COVID Stress Scales” in Greek.

## Methods

### Study design

We conducted a cross-sectional study with 200 participants between November 2021 to February 2022. All participants were adults, and a convenience sample was obtained. We applied the forward-backward translation method to create a Greek version of the “COVID Stress Scales” (Galanis, 2019). First, we conducted a pilot study with 30 participants to assess the face validity of the questionnaire (Galanis, 2013). Minimal corrections were made, resulting in the final version of the “COVID Stress Scales” in Greek. We assessed reliability of the questionnaire with test-retest method in a 10-day window. Also, we assessed validity of the questionnaire with exploratory factor analysis. Data were collected in an anonymous way and study protocol was approved from the Faculty of Nursing, National and Kapodistrian University of Athens (reference number; 370, 02-09-2021).

### Questionnaire

The “COVID Stress Scales” comprised 36 items, while responses ranged from 0 (never) to 4 (almost always). Higher values indicate greater stress. According to the creators of the questionnaire, the “COVID Stress Scales” consist of five factors: (a) danger and contamination fears, (b) fears about economic consequences, (c) xenophobia, (d) compulsive checking and reassurance seeking, and (e) traumatic stress symptoms about COVID-19 (Taylor et al., 2020).

### Statistical analysis

We used exploratory factor analysis to assess the construct validity of the “COVID Stress Scales”. We used the varimax rotation method with an acceptable level for factor loading of 0.4. We set the minimum acceptable value for the eigenvalues at 1. We calculated the Kaiser-Meyer-Olkin measure to assess the adequacy of the sample size considering values >0.8 as acceptable. Also, we performed Bartlett’s test of sphericity considering a p-value <0.05 as acceptable. We assessed reliability of the “COVID Stress Scales” with test-retest method in a 10-day window. In that case, we calculated, Pearson’s correlation coefficient for the 36 items with values >0.7 considered to be exceptional. Also, we calculated Cronbach’s coefficient alpha for the factors that emerged from the exploratory factor analysis with values greater than 0.7 indicate an acceptable level of internal reliability. We used numbers and percentages to present categorical variables and mean, standard deviation, median, minimum value, and maximum value to present continuous variables. We calculated mean scores for each factor. In that case, we added the answers in the items of each factor and then we divided the sum with the total number of items. Thus, each factor had a total score from 0 to 4 with higher values indicate a higher level of stress. All tests of statistical significance were two-tailed, and p-values<0.05 were considered as statistically significant. Statistical analysis was performed with the IBM SPSS 21.0 (IBM Corp. Released 2012. IBM SPSS Statistics for Windows, Version 21.0 Armonk, NY, USA).

## Results

### Demographic characteristics

Detailed demographic characteristics of the participants is shown in Table 1. Most of the participants were females (65%), were married (89%), had children (36%) and had a university degree (73%). Mean age of the participants was 34.1 years old. Among the participants, 26% were diagnosed with COVID-19 and 79% a family member/friend was diagnosed. Twenty-three-point five percent were vaccinated for seasonal influenza and 59.5% were vaccinated for COVID-19.

**Table 1.**
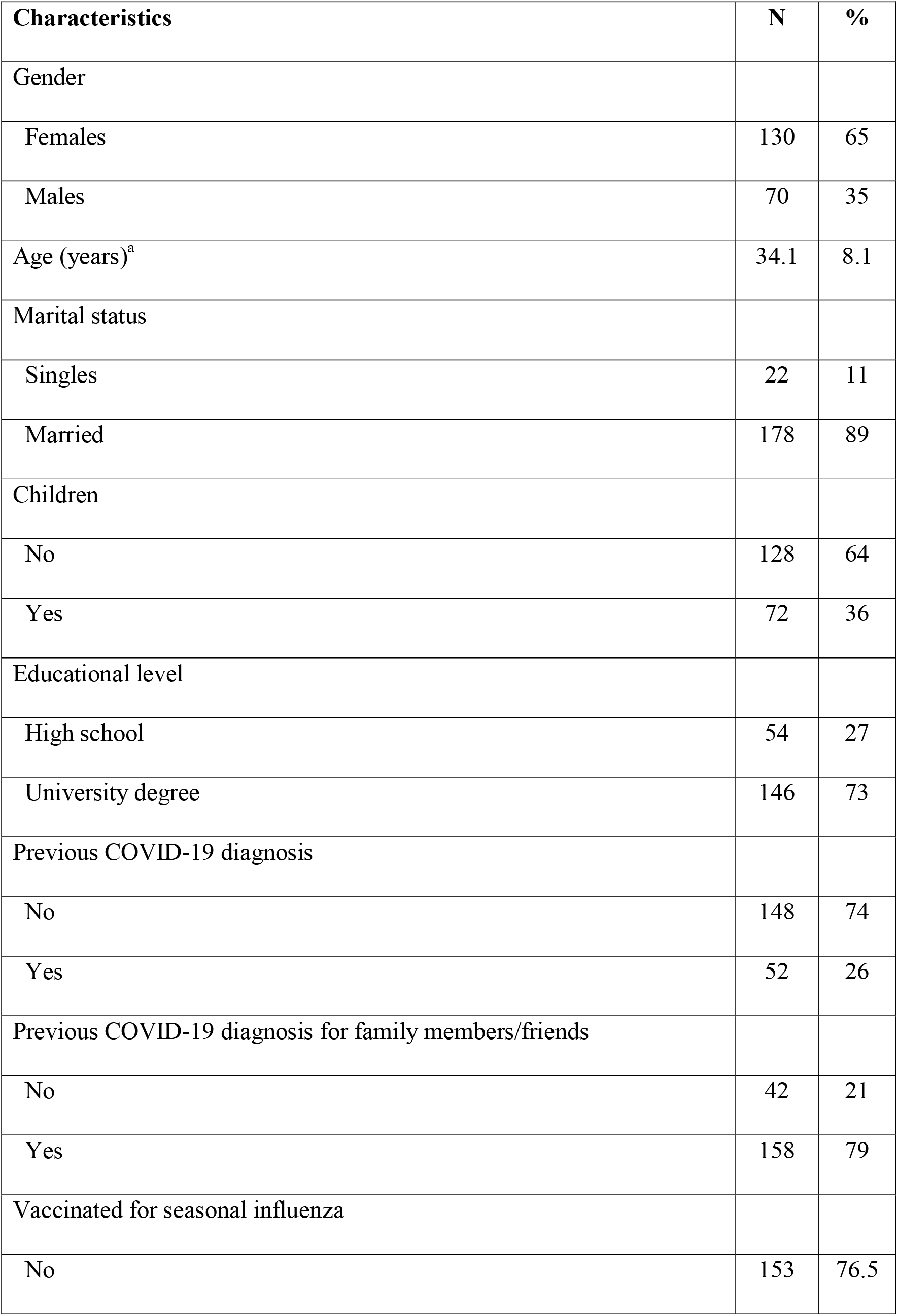
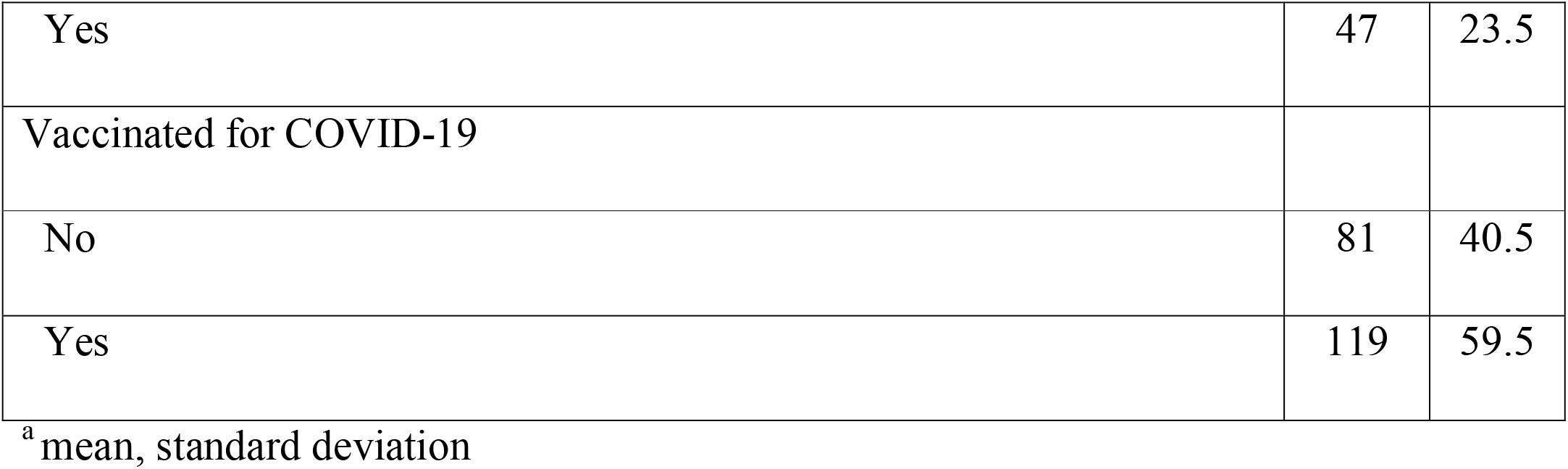
Demographic characteristics of the participants.

| **Characteristics** | **N** | **%** |
| --- | --- | --- |
| Gender |  |  |
| Females | 130 | 65 |
| Males | 70 | 35 |
| Age (years)<sup>a</sup> | 34.1 | 8.1 |
| Marital status |  |  |
| Singles | 22 | 11 |
| Married | 178 | 89 |
| Children |  |  |
| No | 128 | 64 |
| Yes | 72 | 36 |
| Educational level |  |  |
| High school | 54 | 27 |
| University degree | 146 | 73 |
| Previous COVID-19 diagnosis |  |  |
| No | 148 | 74 |
| Yes | 52 | 26 |
| Previous COVID-19 diagnosis for family members/friends |  |  |
| No | 42 | 21 |
| Yes | 158 | 79 |
| Vaccinated for seasonal influenza |  |  |
| No | 153 | 76.5 |
| Yes | 47 | 23.5 |
| Vaccinated for COVID-19 |  |  |
| No | 81 | 40.5 |
| Yes | 119 | 59.5 |

<sup>a</sup> mean, standard deviation

### Factor analysis

Detailed results of the exploratory factor analysis are presented in the Table 2. The p-value for Bartlett’s test of sphericity was <0.001 and the Kaiser–Meyer–Olkin measure was 0.87, indicating that our sample was adequate to perform the exploratory factor analysis. We found five factors including 35 out of items of the “COVID Stress Scales”. Our five-factor model explained 72% of the variance and totally confirmed the factors of the initial “COVID Stress Scales”. In particular, we found the following five factors: (a) COVID-19 danger and contamination (eleven items), (b) COVID-19 socioeconomic consequences (six items), (c) COVID-19 xenophobia (six items), (d) COVID-19 traumatic stress (six items), and (e) COVID-19 compulsive checking (six items). The only item that was not included in our five-factor model was the following: “I am worried that my mail has been contaminated by mail handlers”.

**Table 2.** Exploratory factor analysis for the 36 items of the “COVID Stress Scales”.

| The following questions ask about various kinds of worries that you might have experienced over the past seven days about the coronavirus and the COVID-19 | Factors |  |  |  |  |
| --- | --- | --- | --- | --- | --- |
|  | COVID-19<br>danger and<br>contamination | COVID-19<br>socioeconomic<br>consequences | COVID-19<br>xenophobia | COVID-19<br>traumatic<br>stress | COVID-19<br>compulsive<br>checking |
| I am worried about catching the virus | 0.42 |  |  |  |  |
| I am worried that basic hygiene (e.g., handwashing) is not enough to keep me safe from the virus | 0.67 |  |  |  |  |
| I am worried that our healthcare system is unable to keep me safe from the virus | 0.77 |  |  |  |  |
| I am worried that I can't keep my family safe from the virus | 0.83 |  |  |  |  |
| I am worried that our healthcare system won't be able to protect my loved ones | 0.83 |  |  |  |  |
| I am worried that social distancing is not enough to keep me safe from the virus | 0.73 |  |  |  |  |
| I am worried about grocery stores running out of food |  | 0.84 |  |  |  |
| I am worried about grocery stores running out of cold or flu remedies |  | 0.81 |  |  |  |
| I am worried about pharmacies running out of prescription medicines |  | 0.83 |  |  |  |
| I am worried about grocery stores running out of water |  | 0.91 |  |  |  |
| I am worried about grocery stores running out of cleaning or disinfectant supplies |  | 0.80 |  |  |  |
| I am worried that grocery stores will close down |  | 0.83 |  |  |  |
| I am worried that foreigners are spreading the virus in my country |  |  | 0.83 |  |  |
| If I met a person from a foreign country, I'd be worried that they might have the virus |  |  | 0.87 |  |  |
| I am worried about coming into contact with foreigners because they might have the virus |  |  | 0.88 |  |  |
| I am worried that foreigners are spreading the virus because they're not as clean as we are |  |  | 0.78 |  |  |
| If I went to a restaurant that specialized in foreign foods, I'd be worried about catching the virus |  |  | 0.62 |  |  |
| If I was in an elevator with a group of foreigners, I'd be worried that they're infected with the virus |  |  | 0.68 |  |  |
| I am worried that people around me will infect me with the virus | 0.54 |  |  |  |  |
| I am worried that if I touched something in a public space (e.g., handrail, door handle), I would catch the virus | 0.63 |  |  |  |  |
| I am worried that if someone coughed or sneezed near me, I would catch the virus | 0.75 |  |  |  |  |
| I am worried that I might catch the virus from handling money or using a debit machine | 0.49 |  |  |  |  |
| I am worried about taking change in cash transactions | 0.50 |  |  |  |  |
| I am worried that my mail has been contaminated by mail handlers |  |  |  |  |  |
| I had trouble sleeping because I worried about the virus |  |  |  | 0.78 |  |
| I had bad dreams about the virus |  |  |  | 0.82 |  |
| I thought about the virus when I didn't mean to |  |  |  | 0.82 |  |
| Disturbing mental images about the virus popped into my mind against my will |  |  |  | 0.86 |  |
| had trouble concentrating because I kept thinking about the virus |  |  |  | 0.86 |  |
| Reminders of the virus caused me to have physical reactions, such as sweating or a pounding heart |  |  |  | 0.83 |  |
| Social media posts concerning COVID-19 |  |  |  |  | 0.77 |
| Seeing YouTube videos about COVID-19 |  |  |  |  | 0.67 |
| Seeking reassurance from friends or family about COVID-19 |  |  |  |  | 0.62 |
| Checking your own body for signs of infection (e.g., taking your temperature) |  |  |  |  | 0.57 |
| Asking health professionals (e.g., doctors or pharmacists) for advice about COVID-19 |  |  |  |  | 0.80 |
| Searched the Internet for treatments for COVID-19 |  |  |  |  | 0.89 |
| **Cronbach's alpha coefficient** | 0.93 | 0.94 | 0.94 | 0.93 | 0.89 |

Values express loadings.

### Reliability analysis

Cronbach’s coefficients alpha for the five factors that emerged from the exploratory factor analysis were greater than 0.89 indicating excellent internal reliability. In particular, Cronbach’s coefficient alpha for the factor “COVID-19 danger and contamination” was 0.93, for the factor “COVID-19 socioeconomic consequences” was 0.94, for the factor “COVID-19 xenophobia” was 0.94, for the factor “COVID-19 traumatic stress” was 0.93, and for the factor “COVID-19 compulsive checking” was 0.89. Moreover, Pearson’s correlation coefficients for the 36 items and the five factors were greater than 0.77 (p-value<0.001 in all cases) indicating very good reliability of the “COVID Stress Scales”.

### Descriptive statistics

Descriptive statistics for the five factors of the “COVID Stress Scales” is presented in Table 3. Stress due to COVID-19 danger and contamination was the highest (mean=2.2), and then stress due to COVID-19 xenophobia (mean=1.1), stress due to COVID-19 compulsive checking (mean=0.9), stress due to COVID-19 socioeconomic consequences (mean=0.6), and stress due to COVID-19 traumatic stress (mean=0.5). In general, stress due to COVID-19 was low to moderate.

**Table 3.** Descriptive statistics for the five factors of the “COVID Stress Scales”.

| **Factor** | **Mean** | **Standard**<br>**deviation** | **Median** | **Minimum**<br>**value** | **Maximum**<br>**value** |
| --- | --- | --- | --- | --- | --- |
| COVID-19 danger and contamination | 2.2 | 1.1 | 2.5 | 0 | 4 |
| COVID-19 socioeconomic consequences | 0.6 | 1.0 | 0 | 0 | 4 |
| COVID-19 xenophobia | 1.1 | 1.2 | 0.5 | 0 | 4 |
| COVID-19 traumatic stress | 0.5 | 0.8 | 0 | 0 | 3 |
| COVID-19 compulsive checking | 0.9 | 0.9 | 0.7 | 0 | 3.8 |

## Discussion

We translated and validated the “COVID Stress Scales” in Greek. We found that reliability and validity of the scale was excellent, thus it could be used in Greek samples to measure stress due to the COVID-19. The scale comprised 36 items that are simple and easy to answer them. Our five-factor model totally confirms the five-factor model of the initial “COVID Stress Scales” with the following five factors: (a) danger and contamination, (b) socioeconomic consequences, (c) xenophobia, (d) traumatic stress, and (e) compulsive checking.

One of the factors that we identified through factor analysis was COVID-19 danger and contamination. The COVID-19 pandemic has become a source of fear worldwide and continues to lead people to high levels of fear, anxiety, and insecurity (Heiat et al., 2021; Luo et al., 2021; Muller et al., 2021). Measuring the level of fear, anxiety, and insecurity in different countries and populations could help policy makers to identify high-risk groups in need of public health and education campaigns. For instance, fear is higher in females, in hospital staff, in college students, and in participants in studies in Asia (Luo et al., 2021). Misconceptions and misunderstandings mainly due to media, public awareness level, contradictory data and non-scientific speculations could are the mean reasons for the development of fear and corona-phobia (Heiat et al., 2021).

The second factor that we found in our model was COVID-19 socioeconomic consequences. The COVID-19 pandemic has made many poor countries worldwide face challenging socioeconomic consequences (Buheji et al., 2020; Patterson et al., 2021; Sharma et al., 2021; Singh & Misra, 2020). Lack of vaccines and shortcomings in medical infrastructure are the main reasons that affect negatively the poor societies widening the bridge between the poor and the rich, thus the poor are becoming poorer and the rich are becoming richer. Policymakers should apply immediate control plans and strategies to minimize the impact of the COVID-19 pandemic on the socio-economic activities of the poor. Low-income countries with fewer resources should adopt policies to minimize the impact of the COVID-19 pandemic on the livelihood of the poor.

Also, we found that COVID-19 traumatic stress is another factor in our five-model factor. The prevalence of post-traumatic stress disorder due to the COVID-19 pandemic is high not only to health professionals but also to the general population (Caruso et al., 2021; d’Ettorre et al., 2021; Li et al., 2021; Nagarajan et al., 2022; Sahebi et al., 2021). The prevalence of post-traumatic stress disorder during the COVID-19 pandemic reaches 15% in the general population and 21.5% in healthcare workers. The COVID-19 pandemic has placed people under tremendous psychological stress and prevention of post-traumatic stress disorder is a challenge worldwide. There is a need for urgent appropriate support, interventions to protect people from the psychological impact of the COVID-19 pandemic and healthcare policies to prevent and manage post-traumatic stress disorder.

Finally, we found that COVID-19 compulsive checking is one of the factors that create stress in people. Extreme focus on hygiene measures and contamination during the COVID-19 pandemic could exacerbate obsessive-compulsive symptoms (Cunning & Hodes, 2022). The COVID-19 pandemic appears to be related with a worsening of obsessive-compulsive symptoms especially during the early stages of the pandemic (Guzick et al., 2021; Maye et al., 2022). Moreover, the COVID-19 pandemic has been an enormous stressor for patients with contamination-related obsessive-compulsive disorder. Treatment personalization and effective evidence-based treatment for obsessive-compulsive disorder should be an urgent public health priority.

### Limitations

Our study has several limitations. According to our knowledge, this is the first study that assesses the validity of the “COVID Stress Scales”. We found that reliability and validity of the scale was excellent but further studies in different populations and settings should test the psychometric properties of the scale. Also, our sample comprised 200 participants, thus studies with larger and more representative samples could add more information regarding the “COVID Stress Scales”. More sophisticated analysis such as confirmatory factor analysis could be applied to confirm or not our results.

## Conclusions

We found that the “COVID Stress Scales” is a reliable and valid tool to measure stress due to the COVID-19 in Greek populations. Our five-factor model totally confirms the five-factor model of the initial “COVID Stress Scales” with the following five factors: (a) danger and contamination, (b) socioeconomic consequences, (c) xenophobia, (d) traumatic stress, and (e) compulsive checking. Accurate measurement of individuals’ stress is essential to understand feelings and responses during a major public health threaten as the COVID-19 pandemic. Further studies could expand our evidence and add valuable data regarding the “COVID Stress Scales”.

## Data Availability

All data produced in the present study are available upon reasonable request to the authors

